# Temporal trends in mortality, heart failure hospitalization, and stroke in heart failure patients with and without atrial fibrillation: A nationwide study from 1997-2018 on 152,059 patients

**DOI:** 10.1101/2024.03.14.24304326

**Authors:** Marte Austreim, Nina Nouhravesh, Mariam E. Malik, Noor Abassi, Deewa Zahir, Caroline Hartwell Garred, Camilla F. Andersen, Morten Lock Hansen, Jonas Bjerring Olesen, Emil Fosbøl, Lauge Østergaard, Lars Køber, Morten Schou

**Author notes:** **Address for correspondence:** Marte Austreim, Hjertemedicinsk Forskning 1, Gentofte Hospitalsvej 1, Opgang 6, 3. sal, 2900 Hellerup.

## Abstract

**Background:** Given the many advances in treating heart failure (HF) and atrial fibrillation (AF) separately over the past decades, it remains unclear how the prognosis of patients diagnosed with both conditions has changed over time. We aimed to investigate the temporal trends in clinical outcomes from 1997 to 2018 in patients diagnosed with both HF and AF.

**Methods:** From Danish nationwide registries, we identified 152,059 patients with a first-time HF-diagnosis from 1997 to 2018. Patients were grouped according to year of new-onset HF and AF-status: Prevalent AF (n=34,734), New-onset HF (n=12,691), and No AF (n=104,634). Outcomes of interest were the five-year risk of all-cause mortality, HF-hospitalization, and stroke.

**Results:** Between 1997 and 2018, the proportion of patients with prevalent or new-onset AF increased from 24.7% (n=9256) to 35.8% (n=14,970). The five-year risk of all-cause mortality decreased from 69.1% (95% CI): 67.9%-70.2%) to 51.3% (49.9%-52.7%), 62.3% (60.5%-64.4%) to 43.0% (40.5%-45.5%), and 61.9% (61.3%-62.4%) to 36.7% (35.9%-37.6%) for the prevalent AF, new-onset AF, and no AF group, respectively. Minimal changes were observed in the risk of HF-hospitalization. The five-year risk of stroke decreased from 8.5% (7.8%-9.1%) to 5.0% (4.4%-5.5%) in the prevalent AF group, 8.2% (7.2%-9.2%) to 4.6% (3.7%-5.5%) in the new-onset AF group, and 6.3% (6.1%-6.6%) to 4.9% (4.6%-5.3%) in the no AF group. Simultaneously, patients prescribed anticoagulant therapy within 90 days after HF-onset increased from 42.7% to 93.1% in patients with prevalent AF and 41.9% to 92.5% in patients with new-onset AF.

**Conclusion:** From 1997 to 2018, we observed an increase in patients with HF and coexisting AF. Mortality and stroke risk decreased across all patient groups regardless of AF-status. Anticoagulation therapy increased, and stroke risk in patients with HF and AF was reduced to similar levels as HF patients without AF at the end of the study period.

**Clinical Perspective:** *What is new?:*

- In patients with heart failure, an increasing proportion had concomitant atrial fibrillation in 2018 versus 1997.
- The mortality- and stroke risk decreased for all patients with heart failure over the past two decades, also for patients with coexisting atrial fibrillation, and more patients received guideline-recommended therapy in 2018 versus in 1997.

*What are the clinical implications?:*

- The increasing trend of patients with heart failure and coexisting atrial fibrillation necessitates a better understanding of the coexistence of these two conditions, of which this study provides insight into the evolving prognosis and management landscape of individuals with both heart failure and atrial fibrillation over the past two decades.

**Graphical abstract:** 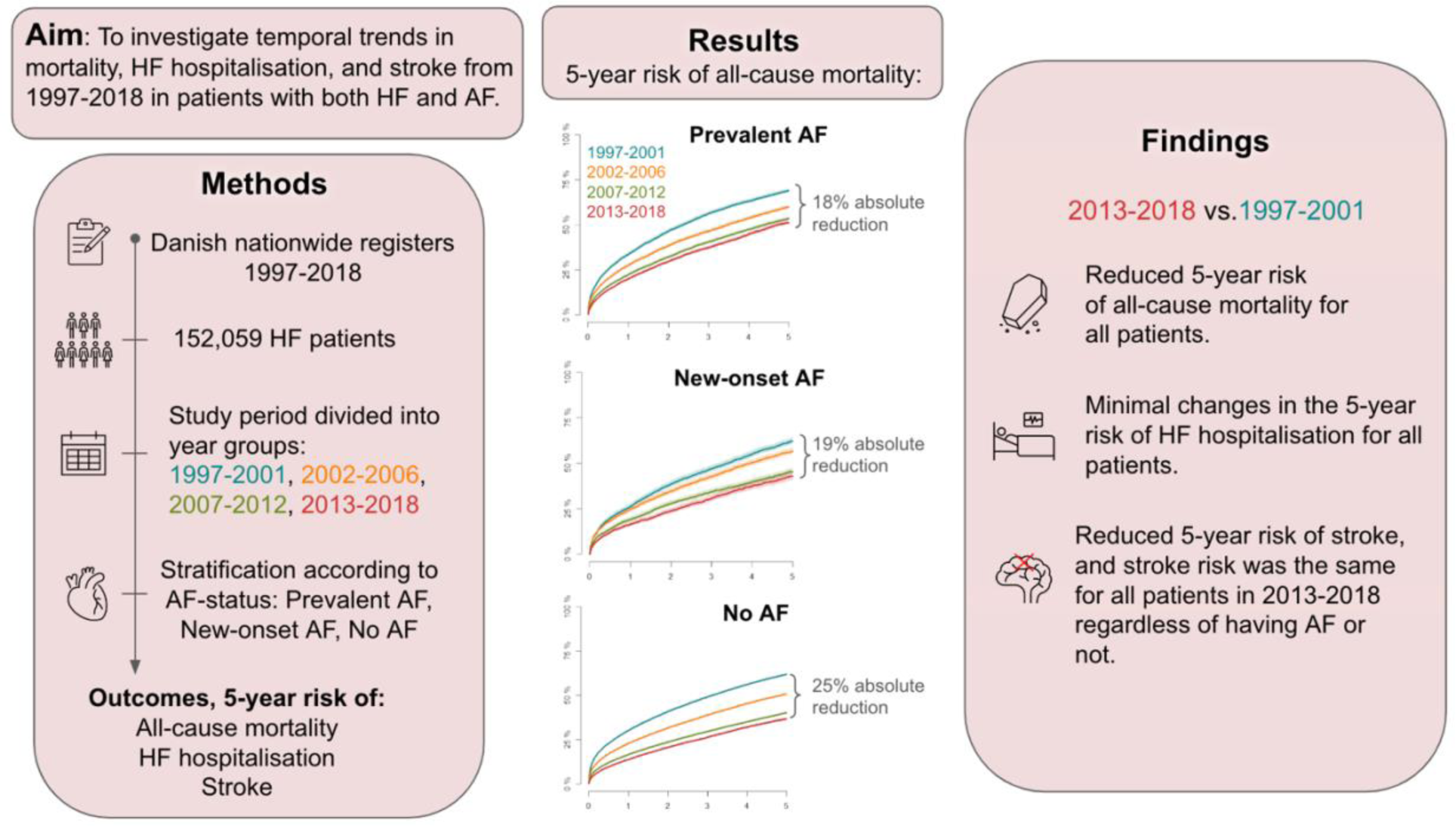

## Introduction

Heart failure (HF) and atrial fibrillation (AF) frequently coexist, exacerbating each other’s clinical manifestations and complicating patient management.^1^ The coexistence of HF and AF is associated with an increased risk of mortality, hospitalization, and stroke.^2^ However, there remains a gap in understanding the evolving prognosis of individuals with both HF and AF over time, particularly given the past decade’s significant advances in the management of each condition. Improvements including renin-angiotensin system inhibitors (RASi), mineralocorticoid receptor antagonists (MRA) and beta-blockers (BB) for patients with HF, and anticoagulation therapy, ablation procedures, and refined risk stratification for stroke in patients with AF, have improved patient prognosis and life expectancy.^3–5^ However, the coexistence of HF and AF poses challenges, leading to divergent opinions among researchers and clinicians regarding optimal management strategies. For instance, while BB have shown mortality benefits in HF with reduced ejection fraction (HFrEF), their efficacy in reducing mortality risk in patients with both HF and AF remains debated.^6^ Conversely, anticoagulation therapy and ablation procedures have shown great promise in mitigating stroke risk and improving outcomes in patients with AF.^7, 8^ Yet, large randomized clinical trials evaluating antithrombotic and anticoagulant treatments have failed in reducing stroke risk in patients with heart failure in sinus rhythm,^9–12^ and the treatment benefits of ablation procedures for patients with both HF and AF remain unclear.^13–15^

Given the past decade’s improvements in managing HF and AF, we aimed to examine the temporal trends from 1997 to 2018 in the five-year risk of all-cause mortality, HF hospitalization, and stroke in patients with coexisting HF and AF. We also examined the prescriptions of BB, MRA, RASi, and oral anticoagulation (OAC, i.e., vitamin K-antagonists and direct-acting oral anticoagulants (DOACs)) within 90 days after receiving the HF diagnosis. By elucidating these trends, our study provides insight into the evolving prognosis and management landscape of individuals with both HF and AF over the past two decades.

## Methods

### Data sources

Data for this study were obtained from Danish nationwide registers. The registers are interconnected through the unique personal identification number assigned to Danish citizens at birth or immigration. The Danish Civil Registration System, The Danish Death Registry, and The Danish National Patient Registry (DNPR) were used to obtain data for analysis and gather baseline characteristics, including sex, date of birth, immigration, emigration, death, diagnosis, and inpatient/outpatient contacts.^16–18^ In DNPR, the diagnoses were classified according to the International Classification of Disease 8^th^ edition (ICD-8) and 10^th^ edition (ICD-10) system. Moreover, The Danish Prescription Registry was used to collect data on redeemed prescriptions from Danish pharmacies using the Anatomical Therapeutic Chemical (ATC) classification system.^19^

### Study population

All patients, 18-90 years of age, presenting with a first-time diagnosis of HF (ICD-10 code I50 or I42) as a primary diagnosis during an inpatient or outpatient contact in the period 1997-2018, were included in this study. The index date was defined as the date on which the patient received the HF diagnosis for the first time. Patients who emigrated before the index date and patients with missing information were excluded from the study. All patients were categorized into three groups based on their AF status. Patients with a primary or secondary AF diagnosis from an in- or outpatient care setting within ten years prior to inclusion were assigned to the “Prevalent AF” group, patients diagnosed with new-onset HF and new-onset AF on the same day were assigned to the “New-onset AF” group, while patients without AF were assigned to the “No AF” group. Further, the different AF status groups were subdivided into four year groups, 1997-2001, 2002-2006, 2007-2012, and 2013-2018, corresponding to the year in which the patient received the HF diagnosis.

### Baseline characteristics

Baseline comorbidities were defined as primary or secondary diagnoses registered within five years before the index date. Concomitant pharmacotherapy was defined as prescriptions redeemed up to 180 days before the index date. Patients with diabetes were defined as either having an ICD-10 code for diabetes (E10-14), a prescription for antidiabetic medication (ATC-code A10), or both in case of receiving a sodium-glucose cotransporter-2 inhibitor (SGLT2-i). Supplemental Table 1 and 2 contain a comprehensive list of the ICD-10 codes and ATC codes used for the baseline characteristics. In addition, we computed the CHA_2_DS_2-_VASc Score (Congestive heart failure = 1 point, hypertension = 1 point, age >_75 years = 2 points, diabetes mellitus = 1 point, stroke = 2 points, vascular disease = 1 point, age 65-74 years = 1 point, sex category (female) = 1 point) for all patients, indicating the presence, or absence of stroke risk (≥ 3 points for females and ≥ 2 points for males). The Charlson Comorbidity Index score was also calculated and presented in Table 1 as counts and percentages of patients with more than three points in the Charlson scoring system.^20^

**Table 1.** Baseline characteristics of patients with HF are stratified according to the year group corresponding to the year of receiving the HF diagnosis.

|  | **1997-2001**<br>(n=37,491) | **2002-2006**<br>(n=34,179) | **2007-2012**<br>(n=38,569) | **2013-2018**<br>(n=41,820) | **Total (n = 152,059)** |
| --- | --- | --- | --- | --- | --- |
| **Age (years), median**<br>**[IQR]** | 76 [67- 82] | 75 [65- 82] | 73 [63- 81] | 73 [64- 80] | 74 [65- 81] |
| **Male, n (%)** | 20,832 (56) | 20,065 (59) | 24,074<br>(62) | 26,241 (63) | 91,212 (60) |
| **CHA2DS2-VASc-score 3**<br>**points or more, female,**<br>**n (%)** | 15,672 (42) | 13,102 (38) | 13,270<br>(34) | 14,129 (34) | 56,173 (37) |
| **CHA2DS2-VASc-score 2**<br>**points or more, male,**<br>**n (%)** | 18,971 (51) | 18,022 (53) | 21,632<br>(56) | 23,406 (56) | 82,031 (54) |
| **Charlson Comorbidity**<br>**Score > 3 points, n (%)** | 2627 (7) | 3663 (11) | 4868 (13) | 5856 (14) | 17,014 (11) |
| **Comorbidities at baseline, n (%)** |  |  |  |  |  |
| **Diabetes** | 5858 (16) | 6164 (18) | 8037 (21) | 9160 (22) | 29,219 (19) |
| **Cancer** | 3168 (9) | 3074 (9) | 4069 (11) | 5482 (13) | 15,793 (10) |
| **Cerebrovascular disease** | 3656 (10) | 3686 (11) | 3999 (10) | 4016 (10) | 15,357 (10) |
| **COPD** | 5887 (16) | 5604 (16) | 5666 (15) | 6154 (15) | 23,311 (15) |
| **Ischemic Heart Disease** | 14,361 (38) | 14,642 (43) | 16,871<br>(44) | 16,046 (38) | 61,920 (41)) |
| **Renal disease** | 1789 (5) | 2231 (7) | 2965 (8) | 3654 (9) | 10,639 (7) |
| **Peripheral vascular**<br>**disease** | 2358 (6) | 2223 (7) | 1917 (5) | 1694 (4) | 8,192 (5) |
| **Pharmacotherapy at baseline, n (%)** |  |  |  |  |  |
| **SGLT2 inhibitor** | 0 (0) | 0 (0) | 0 (0) | 206 (0.5) | 206 (0.1) |
| **Vitamin K-antagonist** | 3705 (10) | 5272 (15) | 7370 (19) | 6144 (15) | 22,491 (15) |
| **DOAC** | 0 (0) | 0 (0) | 268 (1) | 5444 (13) | 5,712 (4) |
| **Beta blocker** | 7067 (19) | 11531 (34) | 16556 (43) | 17672 (42) | 52,826 (35) |
| **MRA** | 3750 (10) | 4514 (13) | 4227 (11) | 3881 (9) | 16,372 (11) |
| **RASi** | 11647 (31) | 14442 (42) | 19160 (50) | 19941 (48) | 65,190 (43) |
| **Digoxin** | 9752 (26) | 5979 (18) | 4575 (12) | 3376 (8) | 23,682 (16) |
| **Cordarone** | 740 (2) | 962 (3) | 1184 (3) | 1065 (3) | 3,951 (3) |
| **Flecainid** | 113 (0.3) | 62 (0.2) | 70 (0.2) | 77 (0.2) | 322 (0.2) |
| **Loop diuretic** | 19953 (53) | 16644 (49) | 16129 (42) | 15145 (36) | 67,871 (45) |
| **Potassium Supplement** | 16578 (44) | 12695 (37) | 12047 (31) | 12088 (29) | 53,408 (35) |
| **Statin** | 2531 (7) | 8101 (24) | 16373 (43) | 17426 (42) | 44,431 (29) |
| **Antidiabetics** | 4649 (12) | 4944 (15) | 6668 (17) | 7789 (19) | 24,050 (16) |
| **Thiazide** | 263 (0.7) | 294 (0.9) | 297 (0.8) | 26 (0.1) | 880 (0.6) |

CHA2DS2-VASc = Congestive heart failure, Hypertension, Age ≥ 75 years, Diabetes mellitus, Stroke, Vascular disease, Age 65-74 years, Sex category (female) COPD= Chronic Obstructive Pulmonary Disease, IQR = Interquartile Range, DOAC = direct oral anticoagulants, MRA = mineralocorticoid receptor antagonists, RASi = renin-angiotensin system inhibitors, including angiotensin-converting enzyme inhibitors and selective angiotensin-1 receptor blockers, SGLT2 inhibitors = sodium-glucose cotransporter-2 inhibitors

### Outcomes and follow-up

The primary outcome of interest was a temporal trend analysis of all-cause mortality, and key secondary outcomes were temporal trend analyses of HF hospitalization and incident stroke from 1997-2018. HF hospitalization was defined as the first overnight hospitalization with HF as a primary diagnosis after the new-onset HF diagnosis. Stroke was defined as receiving an ICD-10 code of DI63-64, DG45, or DI74. The proportion of prescriptions of BB, MRA, RASi, and OAC within 180 days before and 90 days after the index date were also outcomes of interest. Patients were followed until the first event of either an outcome, death, emigration, end of study period (31^st^ December 2018), or completion of the five-year follow-up period.

### Statistical methods

Continuous variables were reported as medians with corresponding interquartile range (IQR), while categorical variables were presented as counts and percentages. A temporal trend analysis using the year groups was computed using the Kaplan-Meier estimator to calculate the five-year cumulative incidence of all-cause mortality for each respective AF status group. The Aalen-Johansen method was used to calculate the five-year cumulative incidence of HF hospitalization and stroke, considering death as a competing risk. Multivariable Cox proportional hazard models were utilized to investigate potential associations between the year groups, with the first year group (1997-2001) serving as the reference. The Cox models were adjusted for age, sex, in- or outpatient HF diagnosis, and relevant comorbidities (ischemic heart disease, chronic kidney disease, chronic obstructive pulmonary disease, cancer, diabetes, cerebrovascular disease, and peripheral vascular disease). The results were presented in a forest plot, exhibiting hazard ratios (HR) and absolute risks (AR), along with their corresponding 95% confidence intervals (CI). Additionally, we calculated the yearly proportion of BB, MRA, RASi, and OAC prescriptions within 180 days before and 90 days after getting the HF diagnosis. Patients who died within the first 90 days after the index date were excluded from this analysis. Data management and statistical analysis were performed using R (version 4.2.1 for Windows, R Foundation for Statistical Computing).^21^

### Supplemental analysis

In addition to the primary and secondary outcomes, we conducted further analyses using the methods described before to calculate the five-year cumulative incidence of developing a bleeding condition, AF hospitalization, and undergoing an ablation procedure. The ablation likelihood was stratified solely by year group, regardless of AF status, since patients with AF after the HF diagnosis may also have been offered ablation years after the HF diagnosis. AF hospitalization was defined as an overnight stay at the hospital with AF as the primary diagnosis. Furthermore, we made a sensitivity analysis using Multivariable Cox proportional hazard models for all-cause mortality, stroke, and bleeding, testing the interaction between year groups and AF status. In this analysis, AF status was defined as patients with a registered AF diagnosis (prevalent AF or new-onset AF) vs. No AF. The analysis was adjusted for the same covariates as the before-mentioned Cox models.

### Ethics

Data for this study were approved for use by Statistics Denmark. According to the Danish regulations regarding observational retrospective register-based studies, ethical approval was not necessary.

## Results

### Patient characteristics

In this study, we included 152,059 patients who received a first-time primary diagnosis of HF between 1997 and 2018, of whom 60.0% were males, and the median age was 74 years. The patient selection is shown in Figure 1. Overall, 34,734 (22.8%) patients belonged to the Prevalent AF group, 12,691 (8.3%) patients to the New-onset AF group, and 104,634 (68.8%) patients to the No AF group. Patients included late in the study period were younger, more burdened with comorbidities, especially diabetes, cancer, and renal disease, and received more medication. Patients with both HF and AF had the highest burden of comorbidities, except for ischemic heart disease, and displayed a greater propensity to receive RASi, BB, and MRA before the index date. While the median age of patients in the No AF group decreased from 75 [IQR: 66-82] to 71 [IQR: 61-79] years, the median age of patients in the prevalent AF group remained consistent throughout the study period at 77 [IQR: 69-83] years. Baseline characteristics stratified according to AF status are provided in Supplemental Table 3.

**Figure 1:**
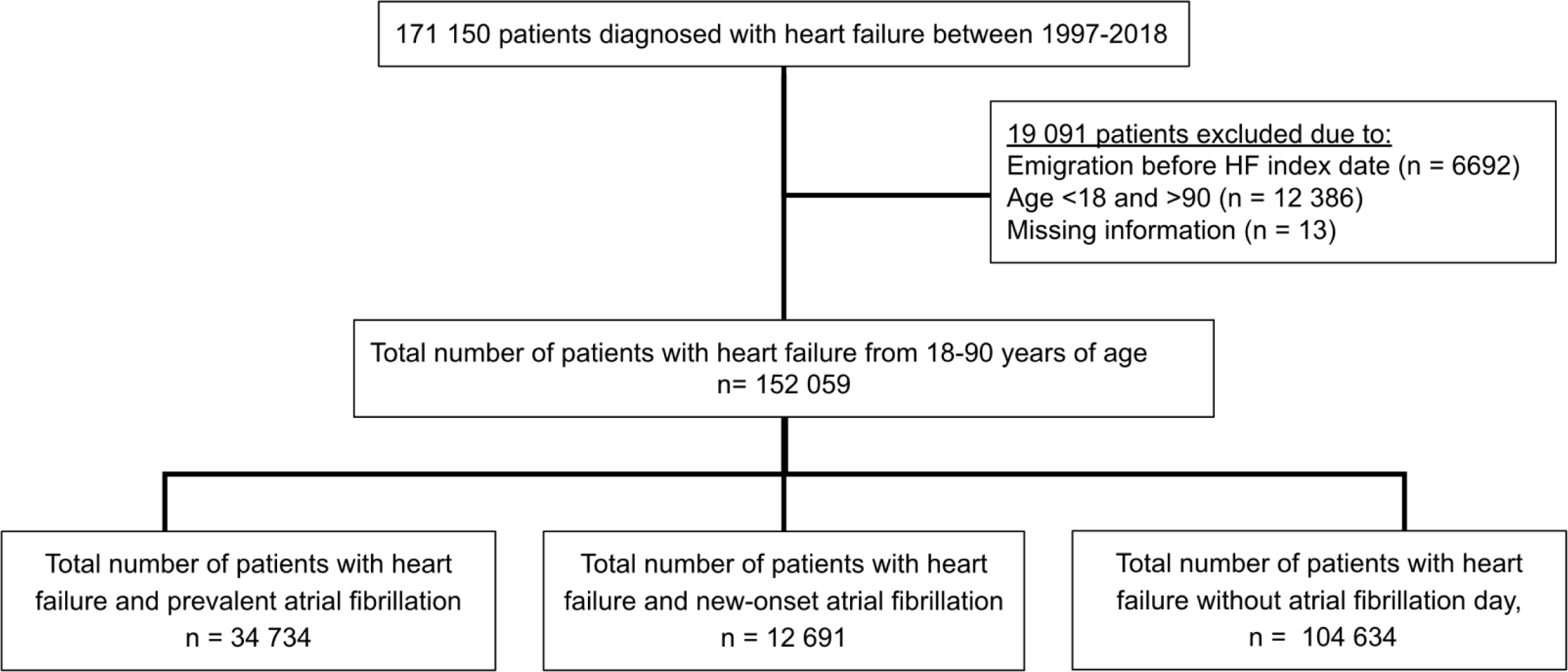
Flowchart of the study cohort. The selection process for patients with HF from 1997 to 2018 is presented as a flow chart, stratified according to AF status. **Abbreviations:** HF: Heart failure, AF: Atrial fibrillation

### Temporal trend over the distribution of patients according to AF status from 1997-2018

Figure 2 illustrates the distribution of patients according to AF status and year group. An absolute increase of 10.7% was seen in the proportion of patients with a prevalent AF between 1997 and 2018 (17.0% to 27.7%). The proportion of patients with new-onset AF remained consistent throughout the study period. The combined percentage of patients in the prevalent AF or new-onset AF group increased from 24.7% to 35.8% between 1997 and 2018 (11.1% absolute increase).

**Figure 2:**
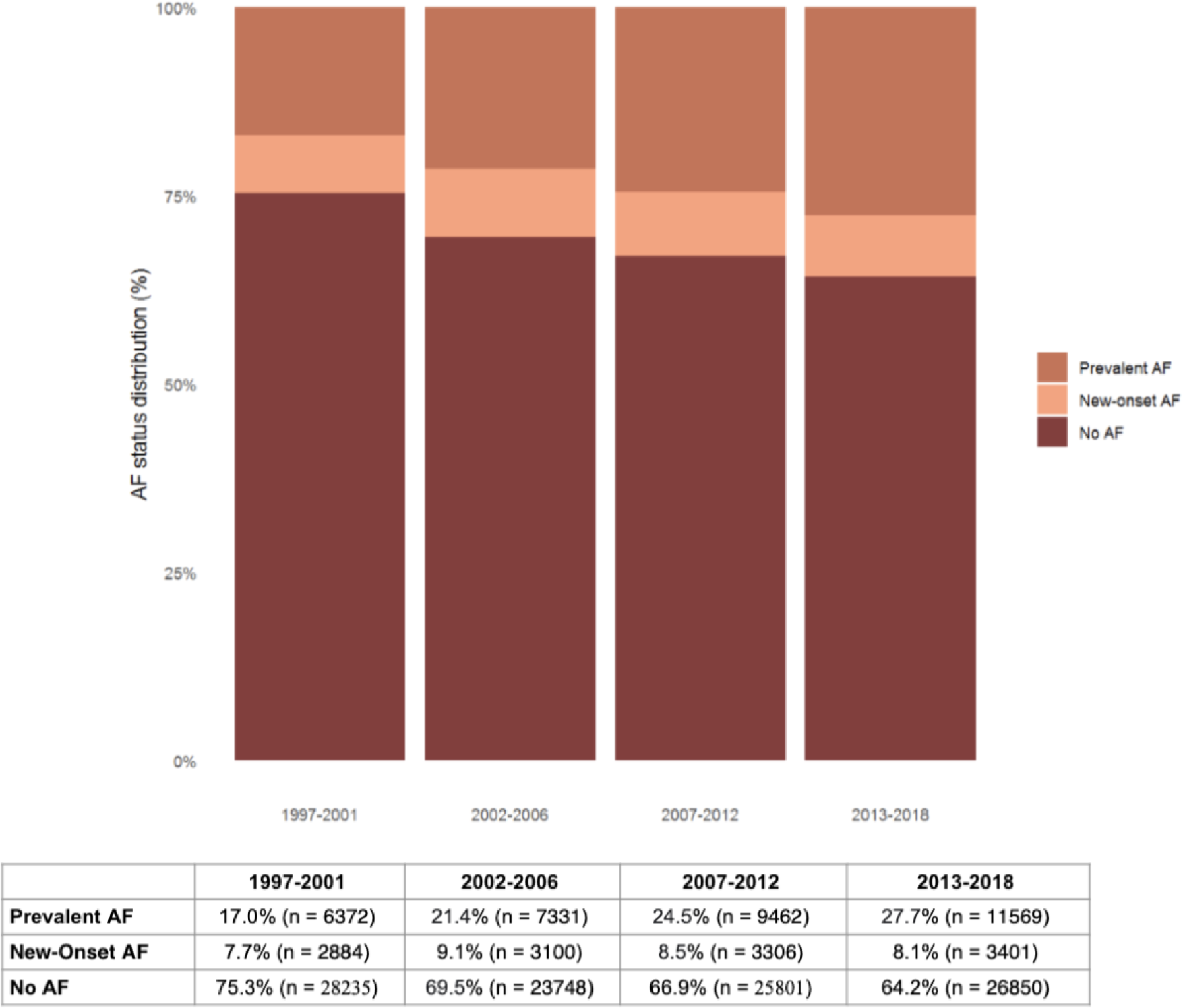
Temporal trends of the distribution of patients with heart failure according to atrial fibrillation status from 1997-2018. Stacked bar chart displaying the distribution of patients with HF according to AF status stratified by year groups: 1997-2001, 2002-2006, 2007-2012, and 2013-2018. **Abbreviations:** HF: Heart failure, AF: Atrial fibrillation

### Temporal trends in all-cause mortality

In Figure 3A, the five-year all-cause mortality risk between 1997 and 2018 decreased from 69.1% (95% CI: 67.9%-70.2%) to 51.3% (95% CI: 49.9%-52.7%) in the Prevalent AF group. In the New-onset HF group, the risk went from 62.3% (95% CI: 60.5%-64.4%) to 43.0% (95% CI: 40.5%-45.5%), and in the No AF group, the risk went from 61.9% (95% CI: 61.3%-62.4%) to 36.7% (95% CI: 35.9%-37.6%).

**Figure 3A-C:**
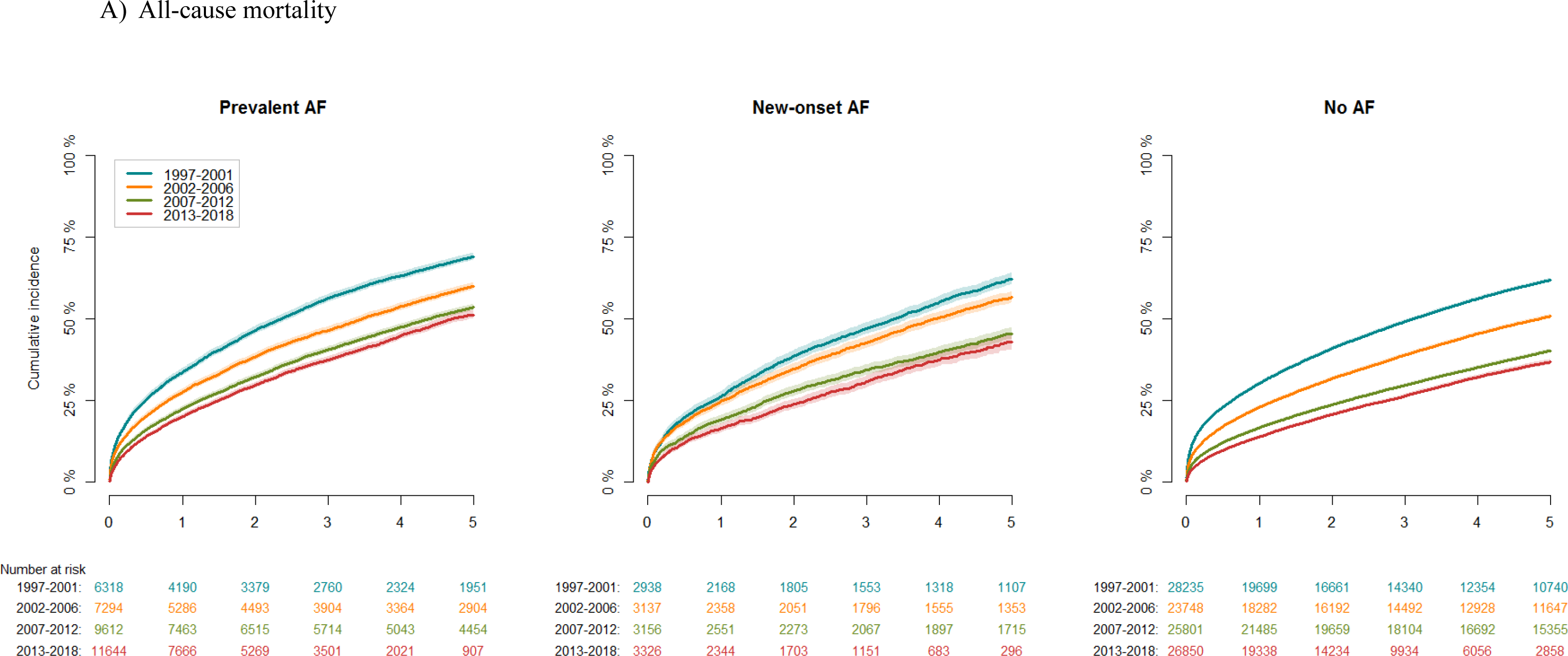
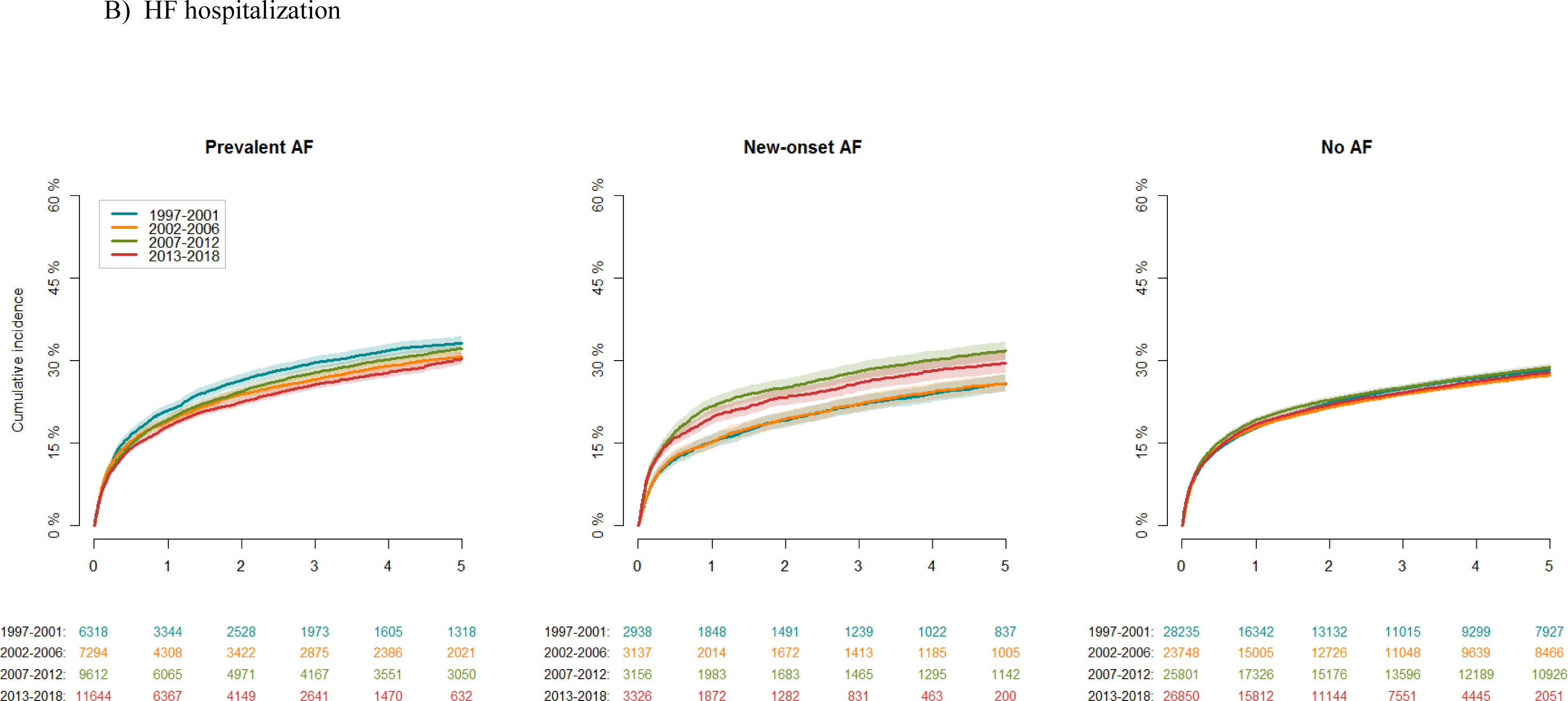
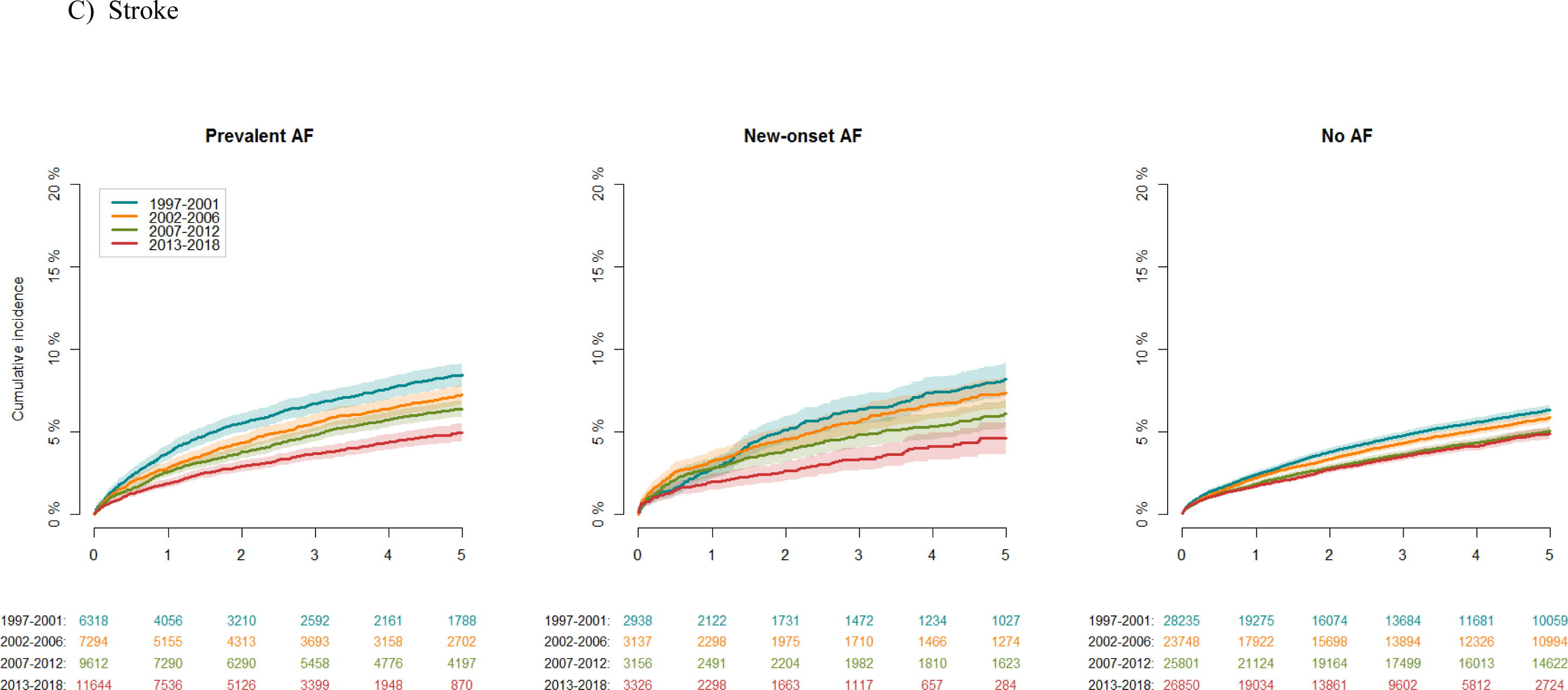
Temporal trends over the five-year cumulative incidence of all-cause mortality, heart failure hospitalization, and stroke stratified by year group and atrial fibrillation status from 1997-2018. The five-year cumulative risk of all-cause mortality, HF hospitalization, and stroke comparing the following year groups: 1997-2001, 2002-2006, 2007-2012, and 2013-2018, in patients with Prevalent AF, New-onset AF, and No AF. **A)** Temporal trends of the five-year cumulative incidence of all-cause mortality. **B)** Temporal trends of the five-year cumulative incidence of experiencing an HF hospitalization event. **C)** Temporal trends of the five-year cumulative incidence of developing stroke. The bands represent 95% CI. Competing risk has been considered while calculating the cumulative incidence of HF hospitalization and stroke. **Abbreviations:** HF: Heart failure, AF: Atrial fibrillation, CI: Confidence intervals.

### Temporal trends in HF hospitalization

The five-year cumulative incidence of HF hospitalization is shown in Figure 3B. For patients in the Prevalent AF group, the risk went from 33.3% (95% CI: 32.1-34.4) to 30.3% (95% CI: 29.2-31.4) between 1997 and 2018. The risk of an HF hospitalization went from 25.8% (95% CI: 24.2-27.4) to 29.6% (95% CI: 27.7-31.5) for patients in the New-onset AF group and from 28.4% (95% CI: 27.8-28.9) to 28.0% (95% CI: 27.2-28.6) for patients in the No AF group over the study period.

### Temporal trends in the risk of stroke

In Figure 3C the five-year cumulative incidence of stroke decreased from 8.5% (95% CI: 7.8%-9.1%) to 5.0% (95% CI: 4.4%-5.5%) for patients in the Prevalent AF group between 1997 and 2018, from 8.2% (95% CI: 7.2%-9.2%) to 4.6% (95% CI: 3.7%-5.5%) for patients in the New-onset AF group, and 6.3% (95% CI: 6.1%-6.6%) to 4.9% (95% CI: 4.6%-5.3%) for patients in the No AF group.

### Prescription of MRA, BB, RASi, and OAC between 1997-2018

Figure 5 illustrates the proportion of patients prescribed MRA, BB, RASi, and OAC within 180 days before and 90 days after the index date. Prescriptions of MRA doubled during the study period, regardless of AF status. The proportion of patients prescribed BB increased from 26.2% to 87.2% in the Prevalent AF group, from 26.8% to 82.6% in the New-onset AF group, and from 20.3% to 75.0% in the No AF group. The initiation of RASi was around 55-60% for all patients in 1997, with a gradual increase to around 75-80% during the study period. The proportion of patients prescribed OAC went from 42.7% to 93.1% in the prevalent AF group, 41.9% to 92.5% for patients diagnosed with New-onset HF, and 9.8% to 17.4% for patients without AF between 1997 and 2018.

**Figure 4:**
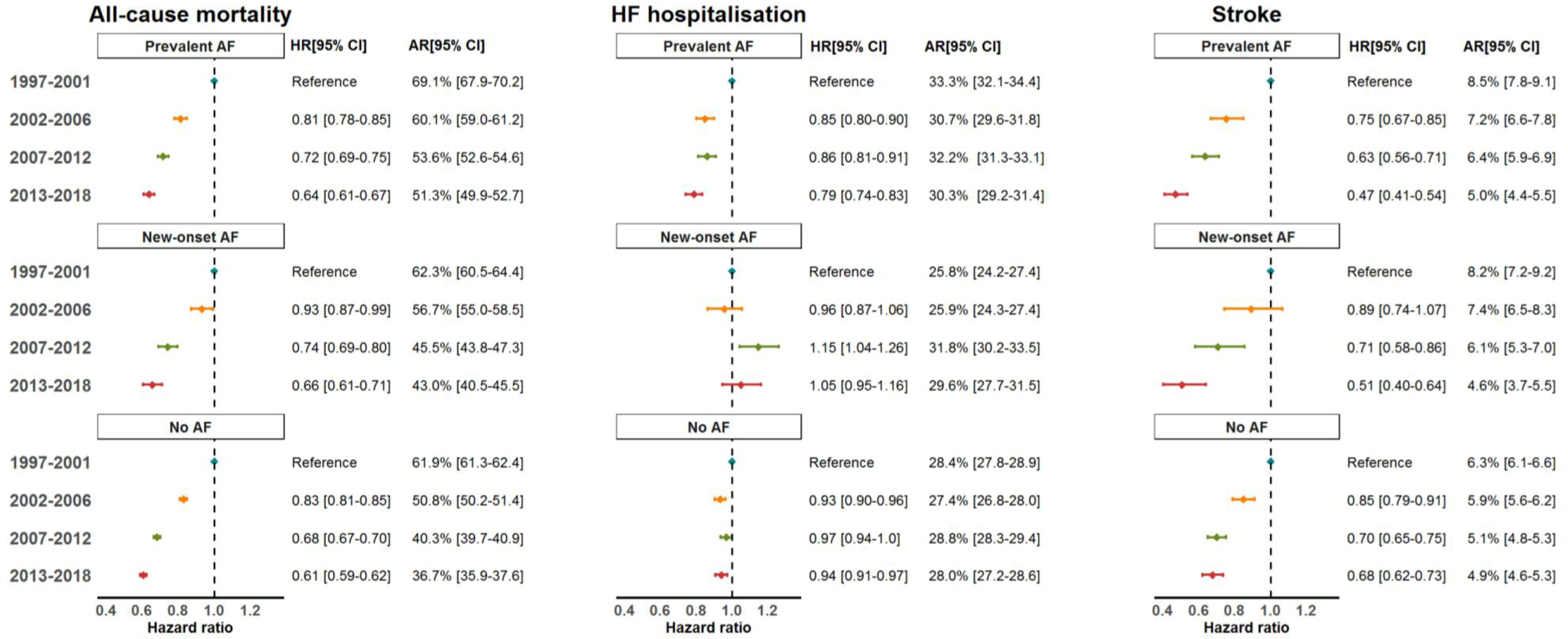
Adjusted Hazard Ratios and absolute risks for five-year all-cause mortality, heart failure hospitalization, and stroke. A forest plot showing hazard ratios and the corresponding 95% CI of five-year all-cause mortality, HF hospitalization, and stroke for the different year groups: 1997-2001, 2002-2006, 2007-2012, and 2013-2018, with 1997-2001 serving as the reference. The model was further stratified according to AF status. The HRs were adjusted for relevant covariates within five years before getting the HF diagnosis (ischemic heart disease, kidney disease, chronic obstructive pulmonary disease, cancer, diabetes, cerebrovascular disease, and peripheral vascular disease), in- or outpatient HF diagnosis, age, and sex. The absolute risk was also calculated and presented as crude risks, with the corresponding 95% CI. **Abbreviations:** HF: Heart failure, AF: Atrial fibrillation, CI: Confidence interval, HR: Hazard ratio, AR: Absolute risk

**Figure 5:**
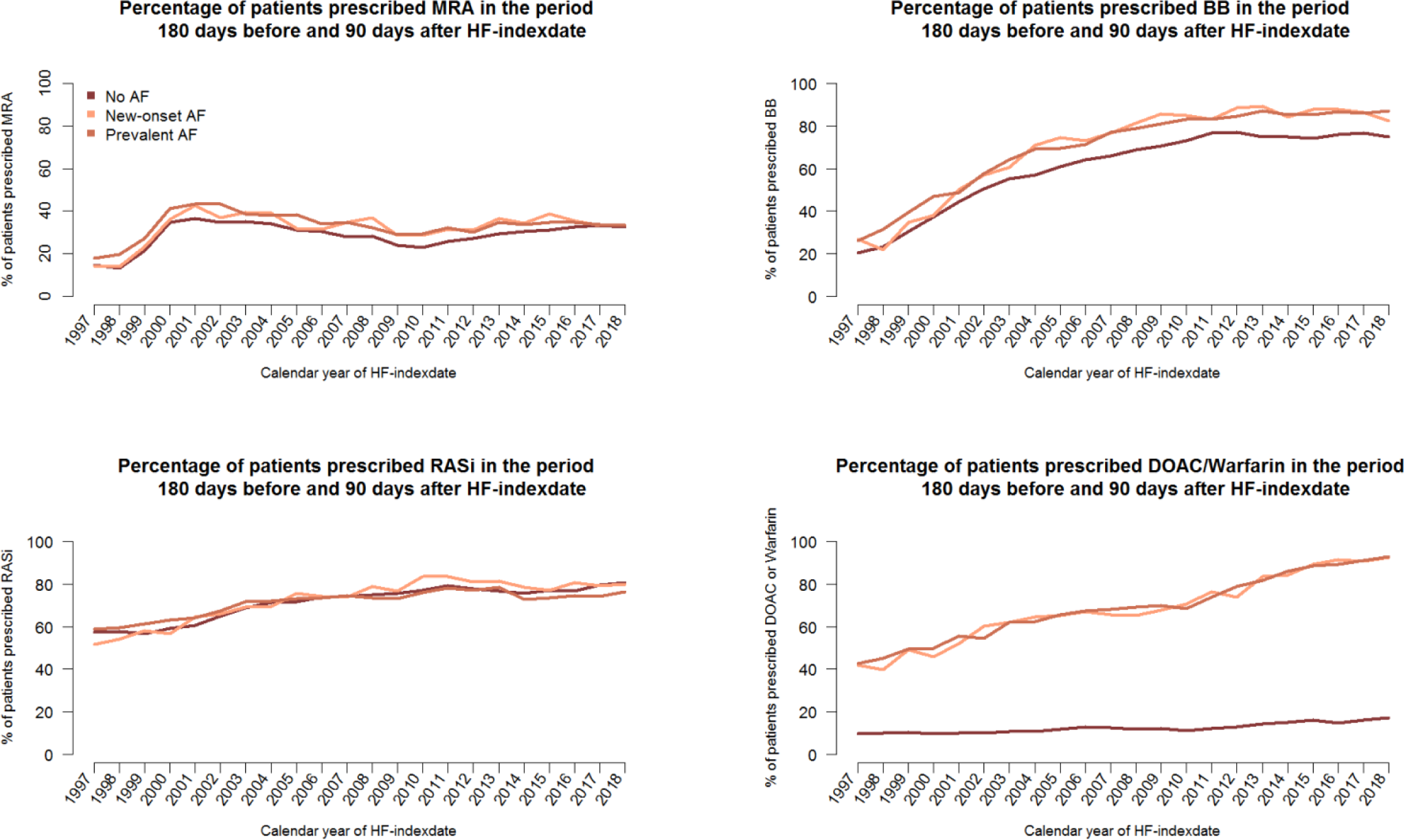
Proportion of patients prescribed MRA, BB, RASi, and OAC in the period 180 days before and 90 days after the index date from 1997-2018. A temporal trend from 1997-2018 showing the proportion of patients prescribed MRA, BB, RASi, and OACs within 180 days before and 90 days after getting the HF diagnosis. Each patient was categorized according to the calendar year in which they were diagnosed with heart failure. The x-axis represents the calendar year, and the y-axis represents the proportion of patients prescribed treatment. **Abbreviations:** HF: Heart failure, AF: Atrial fibrillation, BB: beta-blockers, OACs: Oral anticoagulation therapy, MRA: mineralocorticoid receptor antagonists, RASi: renin-angiotensin system inhibitors

### Supplemental analyses

The temporal trends of the five-year cumulative incidence of developing a bleeding condition stratified by AF status are displayed in Supplemental Figure 2. Patients in the Prevalent AF group had a bleeding risk of 13.4% (95% CI: 12.6%-14.3%) in 1997-2001 and 17.1% (95% CI: 16.2%-18.1%) in 2013-2018. In Supplemental Figure 3, the five-year cumulative incidence of an AF hospitalization went from 17.6% (95% CI: 16.7%-18.6%) to 20.3% (95% CI: 19.3%-21.2%) for patients in the Prevalent AF group, from 20.4% (95% CI: 18.9%-21.8%) to 23.5% (95% CI: 21.7%-25.3%) for patients in the New-onset AF group, and from 4.8% (95% CI: 4.6%-5.1%) to 5.3% (95% CI: 5.0%-5.7%) for patients in the No AF group between 1997 and 2018. We also analyzed the five-year cumulative incidence of undergoing an ablation procedure for the combined AF status groups, which showed an absolute increase of 2.3% from 1997-2001 to 2013-2018 (Supplemental Figure 4). In Supplemental Figure 6, the p-value for interaction between year groups and AF status was < 0.001 for all-cause mortality and stroke and 0.065 for bleeding. Adjusted HR for all-cause mortality was 1.03 (95% CI: 0.99-1.07), 0.88 (95% CI: 0.79-0.99) for stroke, and 1.21 (1.11-1.32) for bleeding in 2013-2018.

## Discussion

In this extensive nationwide cohort study, we investigated the temporal trends of the five-year risk of all-cause mortality, HF hospitalization, and stroke in 152,059 patients with HF and prevalent AF, new-onset AF, or without AF from 1997 to 2018. Three pivotal observations were observed: I) A substantially larger proportion of patients with HF had AF in 2018 vs. 1997, II) The five-year risk of all-cause mortality and stroke decreased from 1997 to 2018, regardless of AF status, III) The risk of stroke was equalized for all patients with HF in 2013-2018 regardless of having AF or not, and an increasing trend of patients with AF were prescribed OACs throughout the study period.

### The prevalence of AF in patients with HF over time

Our study reveals a consistent rise in patients with HF and concomitant AF from 25% to 36% (1997-2018), which aligns with previous non-temporal studies showing a prevalence of 23-69%.^22–24^ The gradual increase in AF prevalence may be attributed to increased focus on AF, lifestyle changes (e.g., increased obesity prevalence), and improved treatment options extending AF patients’ life expectancy.^3, 25^ Notably, the age at which HF was diagnosed in Denmark has not increased, ruling out aging as an explanatory factor. Changes in the HFrEF to HF with preserved ejection fraction (HFpEF) ratio due to more focus on HFpEF and enhanced data collection may further explain our observations.^26, 27^

### Temporal trends in all-cause mortality, stroke, and HF hospitalization

To our knowledge, this study is the first to examine temporal trends over two decades in the risk of all-cause mortality, HF hospitalization, and stroke in patients with HF and AF. The five-year risk of all-cause mortality decreased over time, regardless of AF status (Figure 3A). Patients diagnosed with AF exhibited the highest absolute five-year mortality risk, as well as the highest comorbidity burden and age. Therefore, we made a sensitivity analysis adjusted for age and comorbidities (Supplemental Figure 6A), revealing no significant association between increased risk of all-cause mortality and AF presence in patients with HF. This aligns with findings from other studies.^28, 29^ Advances in HF and AF management, e.g., BB and DOACs, may also have contributed to the decreased mortality risk. Notably, we observed a decrease in mortality even among patients with AF, despite previous studies suggesting no benefit from BB on mortality for these patients.^6^

The risk of stroke also decreased during the study period, possibly due to the enhanced initiation of OACs (Figure 5) and increased focus on stroke prevention.^5, 9^ Consequently, we noted an anticipated rise in the likelihood of developing a bleeding condition (Supplemental Figure 2). While promising outcomes have been reported in previous investigations on ablation procedures.^14, 15^ our findings indicate a limited influence on the observed stroke risk reduction, considering the infrequent utilization of ablation in our study cohort (Supplemental Figure 4). Additionally, an adjusted sensitivity analysis revealed no significant association between AF presence and increased stroke risk (Supplemental Figure 6B).

The risk of HF hospitalization remained consistent throughout the study period for all patients. Denmark’s unique healthcare approach, with outpatient clinics playing a crucial role in managing HF hospitalization events^30^, may have contributed to the low risk of HF hospitalizations compared to other Western countries.^31^ The challenge in clinical practice of distinguishing between hospitalizations for HF or AF in patients with both conditions was addressed by calculating AF admission risk (Supplemental Figure 3). Although no significant decline over time was observed, patients with new-onset AF had a higher risk of AF admission, suggesting a greater cardiac disease burden in this group.

### Temporal trends in HF and AF pharmacotherapy

The proportion of patients prescribed HF and AF pharmacotherapy exhibited an upward trend during the study period (Figure 5), demonstrating an enhanced initiation of guideline-recommended treatment.^3, 4^ Notably, BB exhibited the greatest increase for all patients, followed by OACs for patients with AF. Use of MRA remained low and uniform across all groups, addressing the global underutilization of this important and lifesaving drug.^32, 33^ Finally, the No AF group exhibited a greater reduction in loop diuretic use, suggesting that symptoms and signs of congestion declined most significantly in this patient group (Supplemental Figure 5).

### Strengths and limitations

This study possesses several notable strengths. Firstly, the loss to follow-up was limited. Secondly, selection and inclusion bias were minimized due to the large study population and the use of high-quality data from Danish registers. The ICD-10 codes used in this study have previously been validated with a PPV of 88.0% for the HF diagnosis and 92.6% for the AF diagnosis. Even though the Danish registers provide solid and representative data, certain limitations should be acknowledged. There is a risk of residual and unmeasured confounding due to the lack of clinical observations such as blood pressure, natriuretic peptide levels, left ventricular ejection fraction, body mass index, New York Heart Association Functional class, electrocardiograms (ECG), and information regarding AF subtype (paroxysmal, persistent, or permanent). The lack of access to ECGs also introduced the risk of misclassification of patients.

### Clinical implications

This study addresses the increasing proportion of patients with HF and co-existing AF, which is why this patient group is and will be, of significant relevance for clinicians now and in the future. Our findings also suggest a reduced risk of mortality and stroke from 1997-2001 to 2013-2018, emphasizing the importance of the implementation of neurohormonal blockade and OACs. However, the now equal risk of developing stroke for all patients with HF, regardless of AF status, insinuates a gap in the knowledge of stroke prevention for patients without AF, for whom further studies are needed. Additionally, our findings indicate that patients with AF were older compared to those without AF, suggesting a potential avenue for enhanced prevention of worsening AF early on to mitigate the risk of developing HF later in life.

### Conclusion

This nationwide study on patients with new-onset HF showed an increasing proportion of patients with coexisting AF from 1997 to 2018 and an association between the advancing year group and a decreased five-year risk of all-cause mortality and stroke. Furthermore, the risk of stroke was the same for all patients in 2013-2018, possibly due to the increased initiation of OACs for patients with AF.

## Data Availability

Data for this study were obtained from Dansih Nationwide Registers, and approved for use by Statistics Denmark.

## Non-standard abbreviations and acronyms

HF: Heart failure
AF: atrial fibrillation
RASi: renin-angiotensin system inhibitors
MRA: mineralocorticoid receptor antagonists
BB: beta-blockers
HFrEF: Heart failure with reduced ejection fraction
HFpEF: Heart failure with preserved ejection fraction
DOAC: direct-acting oral anticoagulants
OAC: vitamin K-antagonists and direct-acting oral anticoagulants
DNPR: The Danish Death Registry, and The Danish National Patient Registry
SGLT2-i: sodium-glucose cotransporter-2 inhibitor
ECG: electrocardiogram

## Acknowledgments

None

